# Contacts in context: large-scale setting-specific social mixing matrices from the BBC Pandemic project

**DOI:** 10.1101/2020.02.16.20023754

**Authors:** Petra Klepac, Adam J Kucharski, Andrew JK Conlan, Stephen Kissler, Maria L Tang, Hannah Fry, Julia R Gog

## Abstract

Social mixing patterns are crucial in driving transmission of infectious diseases and informing public health interventions to contain their spread. Age-specific social mixing is often inferred from surveys of self-recorded contacts which by design often have a very limited number of participants. In addition, such surveys are rare, so public health interventions are often evaluated by considering only one such study. Here we report detailed population contact patterns for United Kingdom based self-reported contact data from over 36,000 volunteers that participated in the massive citizen science project BBC Pandemic. The amount of data collected allows us generate fine-scale age-specific population contact matrices by context (home, work, school, other) and type (conversational or physical) of contact that took place. These matrices are highly relevant for informing prevention and control of new outbreaks, and evaluating strategies that reduce the amount of mixing in the population (such as school closures, social distancing, or working from home). In addition, they finally provide the possibility to use multiple sources of social mixing data to evaluate the uncertainty that stems from social mixing when designing public health interventions.

## 1 Introduction

For directly transmitted respiratory pathogens such as influenza, measles and coronaviruses, social mixing patterns shape the risk of individual-level infection [6] and population-level transmission dynamics [29, 18], as well as the effectiveness of control measures targeted at specific age groups [2]. Typically two main approaches have been used to measure social mixing patterns relevant for the spread of disease: inference of contacts based on wearable devices such as proximity sensors [26, 20], or self-recording of contacts [9]. As well as being able to capture age-specific patterns of infection [18], self-recording also allow for details of the type and setting of social contacts, and demographic information about the contacts themselves.

A landmark dataset of self-reported contacts was the POLYMOD study [22], which collected social mixing data for 7,290 participants across eight European countries. These data have been widely used to understand the epidemiology of infectious diseases [25] and inform policy-relevant disease modelling [23, 2]. However, the sample size for each country (e.g. 1,012 participants for Great Britain) limit the ability to stratify by multiple demographic factors and still obtain precise estimates of social mixing within those groups, and does not have details about the location of participants, which meant social contacts could not be compared between spatial covariates such as urban and rural setting. Moreover, the POLYMOD study was conducted in 2005-6, and so patterns may have changed since then.

To generate a more contemporary large-scale dataset on social mixing and movement patterns in the United Kingdom, the BBC Pandemic project recruited over 86,000 participants between September 2017 and December 2018 as part of a public science project linked to a BBC4 documentary [17]. Here we present high resolution age-specific social mixing matrices based on data from over 40,000 participants, stratified by key characteristics such as contact type and setting.

## 2 Data collection

There were two components to the BBC Pandemic study, one focused on the town of Haslemere [16], and another focused on the wider UK population [17]. Here we present data from the UK national study. Upon using BBC Pandemic app to join this study, users first entered their basic demographic information, including age, household size, gender and occupation. The app then recorded their approximate location at hourly intervals for a 24 hour period. At the end of this period, users recorded each social contact they had made during this period, including information on: the contact’s age; the type of interaction (conversational contact, defined as face-to-face conversation of three or more words, or physical contact); the setting of the contact (home, work, school, other); and whether the participant had spoken to that person before.

Overall, over 86,000 participants started the survey and filled out their profile. Participants with no encounter or location data were excluded, as were users whose location recordings were all outside the UK. This leaves a rich dataset of around 55,000 participants. Of those, 40,177 completed the study and reported their social contacts at the end of the survey – this is the focus for this paper.

The data collection process had some limitations. In particular, the initial version of the app had the default age of a contact as 50-years-old. Participants were free to change the value on slider, but if they just clicked through, it would record that contact’s age as 50. As a result, the early data had more contacts of this age than was plausible. In our analysis, we therefore remove users with 3 or more contacts of the age exactly 50 (4,007 users dropped). These users together reported 101,880 contacts out of which 24,171 were aged exactly 50-years-old. Second, the initial version did not allow users to record precisely zero contacts: these users may thus be missing from our denominators, we do not expect this effect to be large and for simplicity have not attempted to correct for it here.

### 2.1 Ethical considerations

Information was provided and consent obtained from all participants in the study before the app recorded any data. The study was approved by London School of Hygiene & Tropical Medicine Observational Research Ethics Committee (ref 14400).

## 3 Methods

We follow [29] to infer mixing matrices from the self-reported contact data. We group the study participants and their contacts by age into following age groups: 0-4, 5-9, 10-12, 13-14, 15-17, 18-19, 20-21, 22-24, then 5 year age bands from 25 to 74, with a single category for those aged 75 and over: the finer structure chosen to provide higher resolution for school and university ages. For each of those age groups, we find *t*_*ij*_: the total number of reported contacts over the course of 24 hours between participants in age group *j* and their reported contacts of estimated age group *i*. To find the mean number of contacts (*m*_*ij*_) who are in age group *i* as reported by participants in age group *j* we divide *t*_*ij*_ by *n*_*j*_ – the total number of participants in age group *j*. This results in the ‘social contact matrix’ **M** = (*m*_*ij*_), where *m*_*ij*_ = *t*_*ij*_*/n*_*j*_. This is the *raw contact matrix* as derived from our study data.

We can deduce more from our reported contacts as the contacts are reciprocal (if person A was in contact with B, that means that the person B was also in contact with person A). On a population level that means that the total number of contacts from age group *j* to *i*, must be equal to the total number of contacts from age group *i* to age group *j*. As the sample of participants might have a different population structure than the wider population, this step depends on the country-specific population structure: we define *w*_*i*_ as the total population size of the age group *i*. In our case, volunteers needed to be in the UK to participate in the study, so we use the 2018 mid-year estimate of the population structure of the UK (available from ONS [1]). The reciprocal matrix **C** = (*c*_*ij*_) gives the *population contact matrix*, where 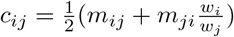 [29].

The population matrix **C** is of particular importance for infectious disease dynamics because it is related to the next generation matrix [7, 27]. The next generation matrix **N** captures how the infection spreads when pathogen is first introduced in a population, and its (*i, j*) entry gives the expected number of new infections in compartment *i* produced by in infected individual introduced into compartment *j*. As a result, the dominant eigenvalue of **N** is equal to the basic reproduction number *R*_0_ or the expected number of secondary infections caused by a single individual introduced to a completely susceptible population. The relationship between the two matrices is 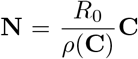 where *ρ*(**C**) is the dominant eigenvalue of **C** (its spectral radius). Analogous to the eigenvector representing stage-specific contributions to overall population reproduction in demographic theory [3], the dominant eigenvector here gives an indication of which age-groups most contribute to transmission in the population, assuming no age-specific differences in susceptibility or infectiousness (Fig 2).

**Figure 1:**
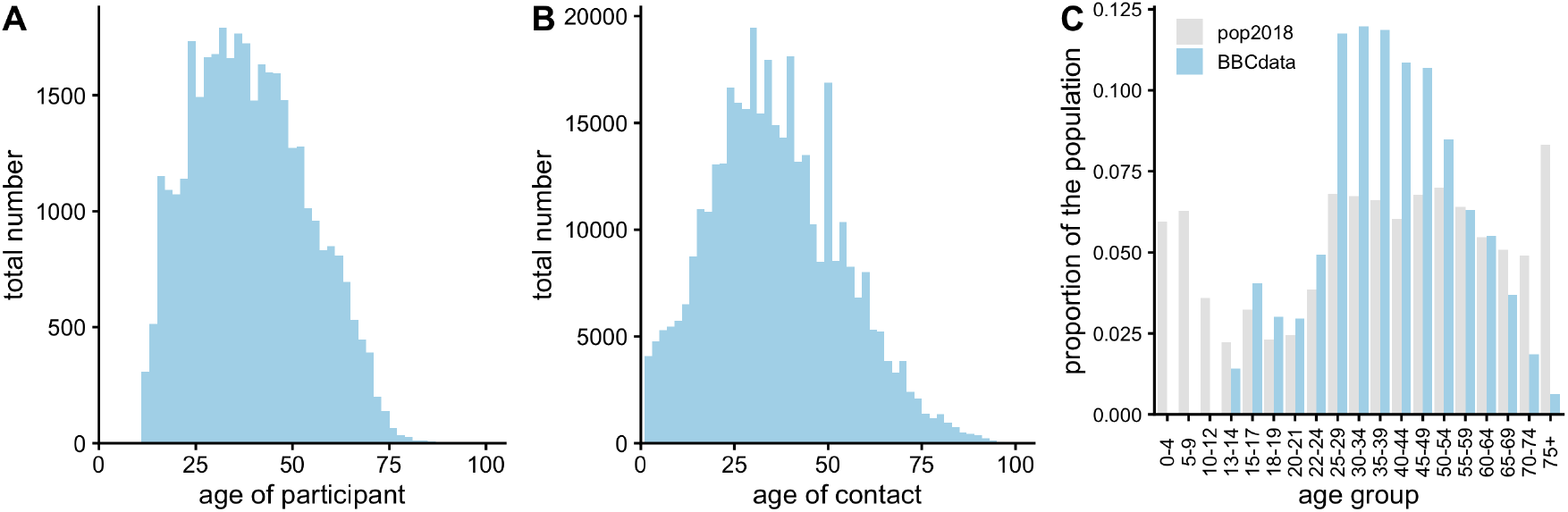
(A) Age distribution of participants included in this study, (B) distribution of ages of contacts, (C) population structure of the study population compared to UK mid-year estimate for 2018 (ONS data [1]).

**Figure 2:**
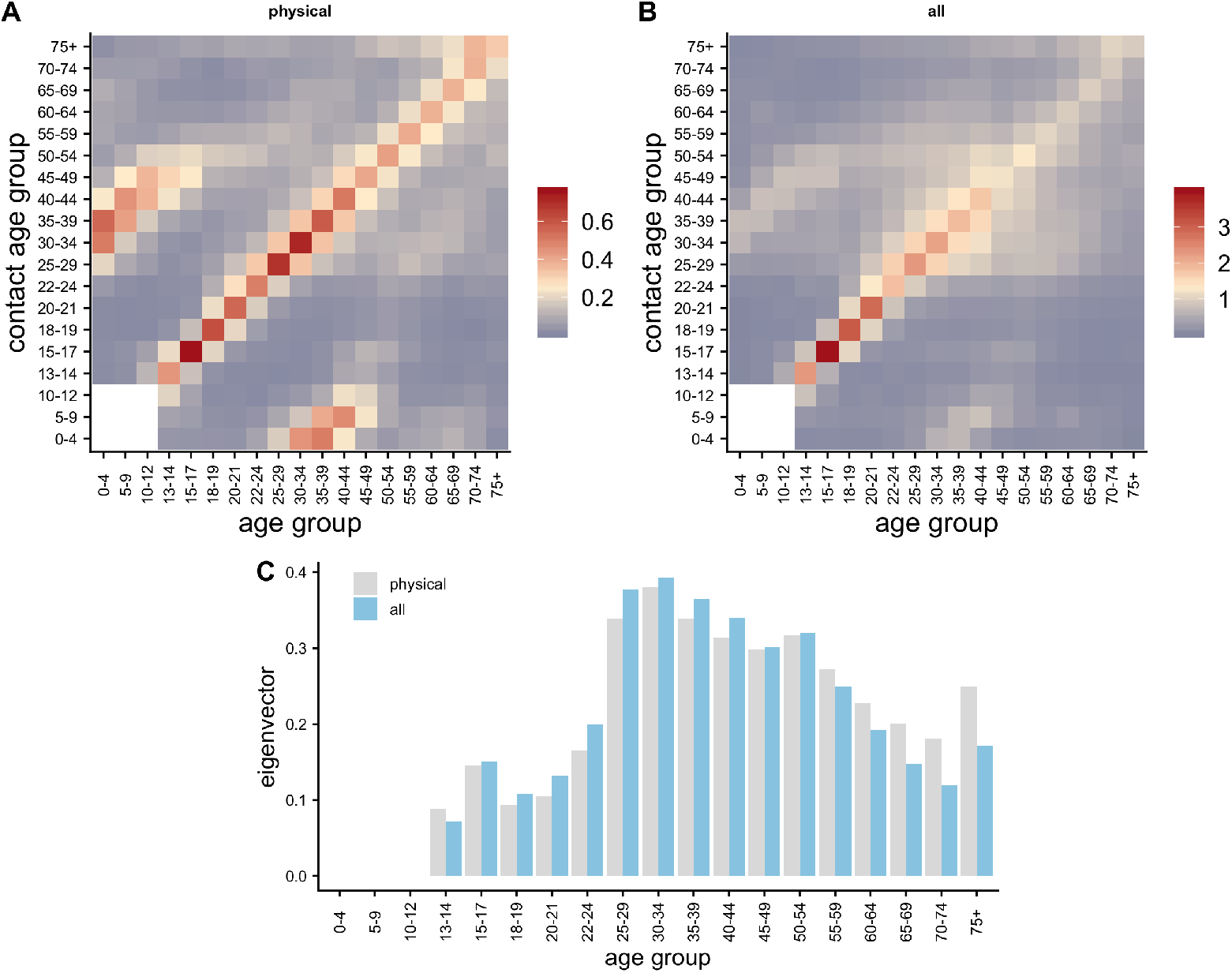
(A) Population contact matrix capturing all conversational contacts, (B) population contact matrix inferred from all physical contacts in the study, with white indicating missing values, (C) dominant eigenvectors of the symmetric subset of the two matrices without missing values.

We repeated these calculations stratified by type of contact (physical or conversational) and by context (home, work, school, other), resulting in 8 matrices each for the *raw contact matrix* shown in Fig S1 and *population contact matrix* in Fig 3. We further stratify these matrices by contacts made during the week (Fig S2) and during the weekend (Fig S3). As participants filled out the contact survey at the end of the 24 hour period, we define the weekend based on the time they activated the app in the following way. To avoid weekend/week overlap, we consider participants who activated the app after 18:00 on Fridays and before noon on Sundays as those reporting their weekend contacts, and those who activated the app after 18:00 on Sundays and before noon on Fridays as those reporting contacts made during the week. This excludes 2,451 users with 24,983 contacts that report their contacts both during the week and the weekend.

**Figure 3:**
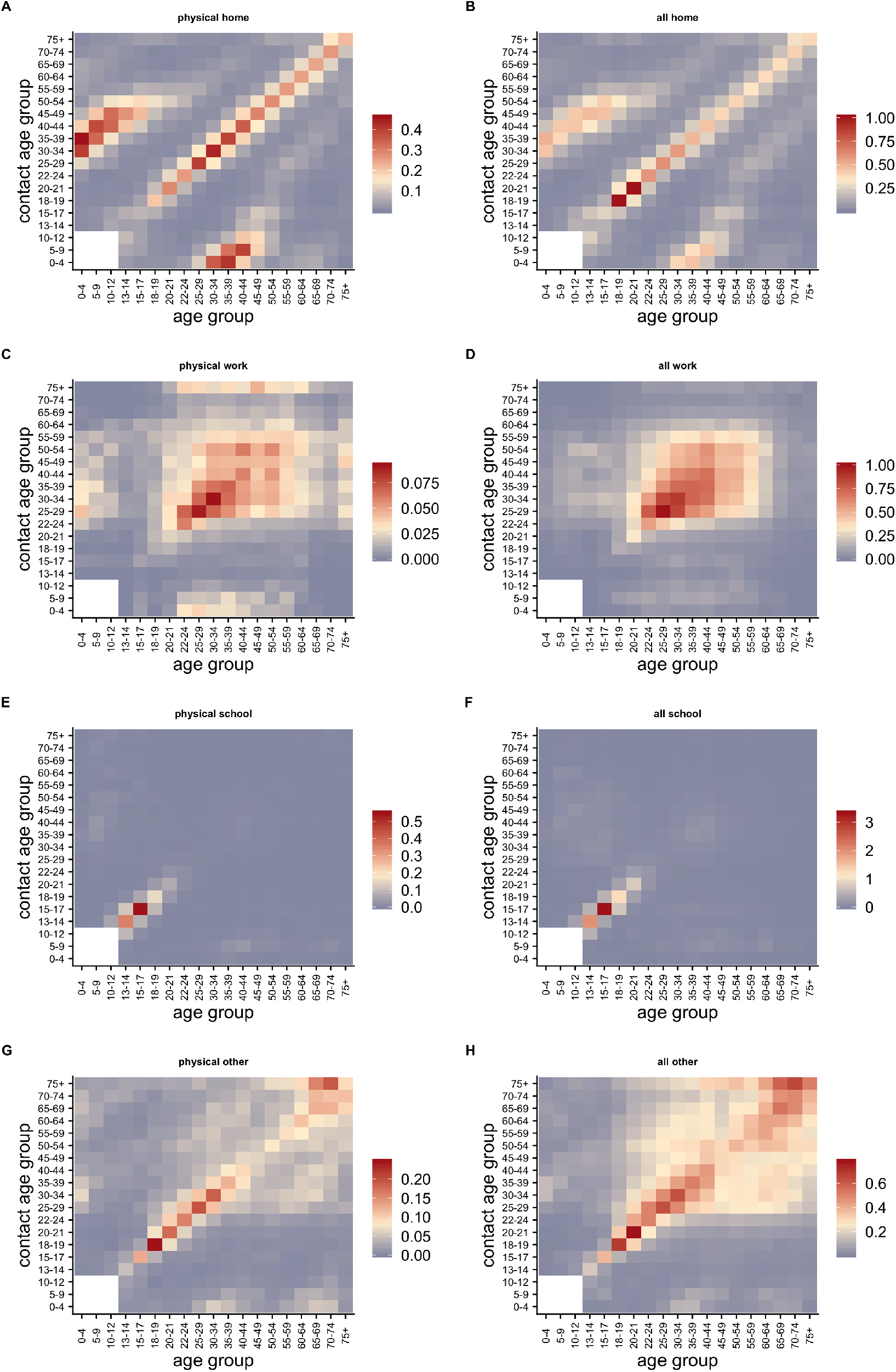
Population contact matrices. Mean number of contacts reported by participants of given age groups adjusted for reciprocity of contacts, **C**, assuming population age structure given by the 2018 mid-year estimate from ONS [1]. Matrices are by different encounter type (physical only or all contact, in respective columns) and by different encounter context (home, work, school or other in respective rows).

We also analysed the relationship between the density locations participants typically visited and their social contacts. We focused on a subset of the data with participants that had at least 10 recorded GPS locations (n=26,153). We used data from 2011 Lower Layer Super Output Areas for England and Wales, Small Areas for Northern Ireland, Data Zones for Scotland to estimate the population density per square km for each recorded GPS location in our dataset, with density layers matched to GPS location based on whichever had the nearest centroid. We then calculated the mean density across all GPS locations recorded for each participants.

Finally, we compared reported household size with social contacts for participants that had at least one reported physical or conversational contact (n=40,575). To explore the relationship between household size, average density of GPS tracks and social contacts, we used a generalized additive model [30]. The model was defined as *g*(*E*(*y*)) = *b* + *f* (*x*) + *a*, where *y* was the binary outcome variable (i.e. reported contacts made), *x* was the predictor (i.e. household size or average density of GPS tracks on *log*_10_ scale), *g* was the link function, *b* was the intercept, *a* was age (to adjust for possible confounding), and *f* was a smooth function represented by a penalized regression spline. Results for fitted GAMs are shown for *a* = 30.

## 4 Results

The main dataset used in this study consisted of 36,155 participants reporting 378,559 contacts. For ethical reasons, participants in the BBC study had to be at least 13 years of age, although the number of younger participants tails off gradually rather than a hard cutoff at this age (Fig 1A). Total reported contacts (i.e. the sum of physical and conversational contacts) were distributed across a wide range of age groups, with a peak reflecting the peak in age of participants, and spikes likely representing bias to chose round numbers as estimated ages of casual contacts. The participant population in the study under-sampled the youngest and oldest groups relative the underlying UK population (Fig 1C).

The total conversational and physical contacts varied greatly across different age groups (Fig 2A,B). On average, participants reported over three times more conversational contacts than physical contacts, and contacts that spanned a larger age range. The very strong diagonal density of contacts (Fig 2B) is characteristic of strong age-assortative mixing, and the sub-diagonal density captures interactions between children and their parents. The dominant eigenvectors of the matrices, which indicate the age groups that would drive transmission during the exponential phase of an epidemic simulated using these data, are highest for the 30-34 age group (Fig 2C). In general, eigenvectors based on physical only contacts and all contacts (both conversational and physical) are very similar except for the ages over 65 where relative dominance is higher for physical contacts.

The measured age-specific social mixing matrices also varied considerably between different types and settings. Reported contacts at home in the *population contact matrix* followed a strong age-assortative pattern (strong diagonal band), with inter-generational mixing shown by the offdiagonal bands in contacts (Fig 3A-B), which is especially pronounced in physical contacts at home. Contacts at work showed less age-assortativity than contact at home (Fig 3C-D), and were predominantly non-physical (Fig 3D). Within school-aged groups, more contacts were reported on average at school than in other settings (Fig 3E-F), but for a very narrow age-band. Overall contacts in other settings (i.e. not home, work or school) were age assortative for younger groups, but less assortative for older groups, with an off diagonal peak in contact intensity between older participants and other adults (Fig 3G-H). Physical contacts in other settings were less common, but also exhibited the transition from age-assortativity to less structured mixing in older age groups (Fig 3G).

Stratifying these contacts by type and context further by those made during the week versus during the weekend shows temporal changes in the average number of contacts in different settings (Fig 4). Both physical and all contacts are higher at home and at ‘other’ locations (not home, school or work) during the weekend, while there is marked decrease in contacts at work and at school, as might be expected.

**Figure 4:**
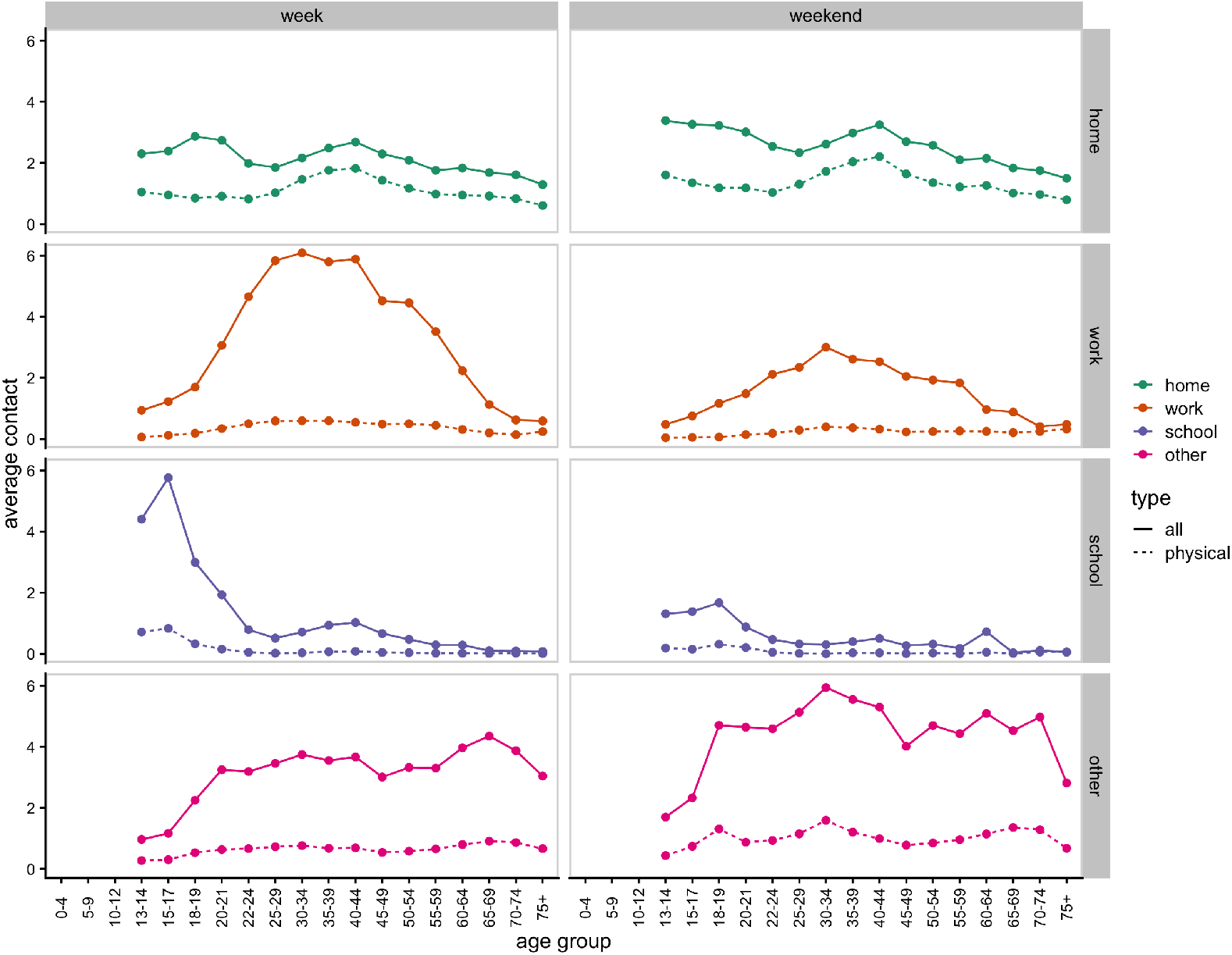
Mean number of contacts for an average person (assuming 2018 population structure from ONS) during the week and during the weekend by encounter context (home, work, school or other, in respective rows) and type of contact (physical only shown with dashed lines or all contacts in solid line).

**Figure 5:**
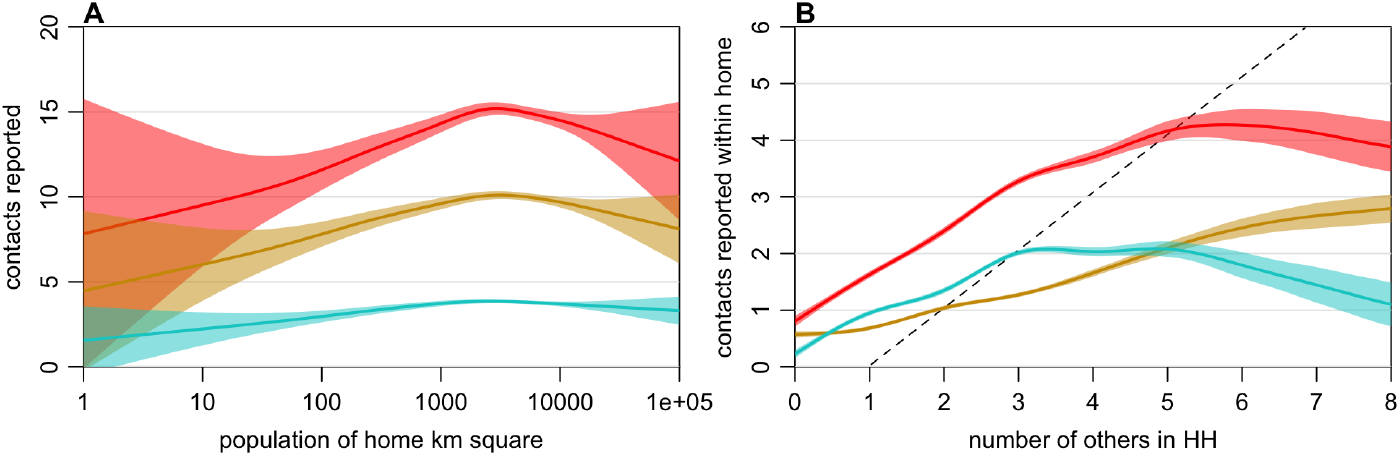
Relationship between density of locations visited, household size and reported contacts. (A) generalized additive model (GAM) fits of average density of locations visited and reported total contacts (red), conversational contacts (gold) and physical contacts (cyan), adjusting for age. (B) GAM fits of number of household occupants (not including participants) and total contacts within the home (red), conversational contacts within the home (gold) and physical contacts within the home (cyan), adjusting for age (results shown for 30 year olds).

We found that participants who typically had recorded GPS tracks in lower density areas had fewer contacts than those that spent most of their time in more dense areas (5A). Participants who spent most time in areas with density between 1000 and 10,000 people per km reported 15 total contacts on average, whereas participants in areas with fewer than 10 people per km recorded fewer than 10 contacts on average. There was a positive association between contacts within the home and household size for participants who lived with up to 3 additional people (5B). However, conversational contacts within the home saturated at around 4 in households of size 5 or larger, and physical contacts saturated at 2 in households of size 4–6, before declining again. We also found many contacts reported on average within the home for the 10% of participants who lived alone (self-reported household size of 1): although 62% of participants who lived alone reported no contacts within the home, the remaining 38% of these participants reported 2.4 contacts within the home on average.

## 5 Comparison with POLYMOD data

To date, the gold-standard for modelling age-specific contact patterns in many settings has been the POLYMOD dataset collected in 2005/06 [22]. In Great Britain, the POLYMOD study had 1,012 participants reporting a total of 11,876 contacts (11.74 contacts on average). This is higher than the daily average reported number of contacts in the BBC contact dataset with 36,155 participants reporting 378,599 contacts (10.47 contacts reported on average). For most age groups, the total number of contacts per day for an average person (assuming 2018 population structure [1]) is remarkably similar between BBC and POLYMOD datasets, especially for ages over 60 (Fig 6A). In BBC contact data a smaller proportion of those contacts are physical (and correspondingly a higher proportion of contacts are conversational only). The reduction in the average number of contacts for ages 10-19 is striking between POLYMOD and BBC datasets. While for the 10-14 age group this may be affected with the fact that we are only sampling a subset of this age group and assuming that 13-and 14-year-olds are representative of the entire age group, for 15- to 19-year-olds this reduction is real. This could be the effect of our sampled population, or it might be a signature of a real change in social contacts of teenagers since POLYMOD. A survey of over 1,000 13- to 17-year-olds in the US in the 2018 showed that compared to 2012 teens’ preference for direct face-to-face communication with friends has declined substantially (from a half to a third) while the interactions over social media have increased [24]. A similar trend in digitisation of teenager interactions is likely to be present also in the rest of the world with comparable teenage mobile phone use, and it is possible that the reduction in both conversational and physical contacts in our dataset is a reflection of teenage interactions moving away from face-to-face to social media.

**Figure 6:**
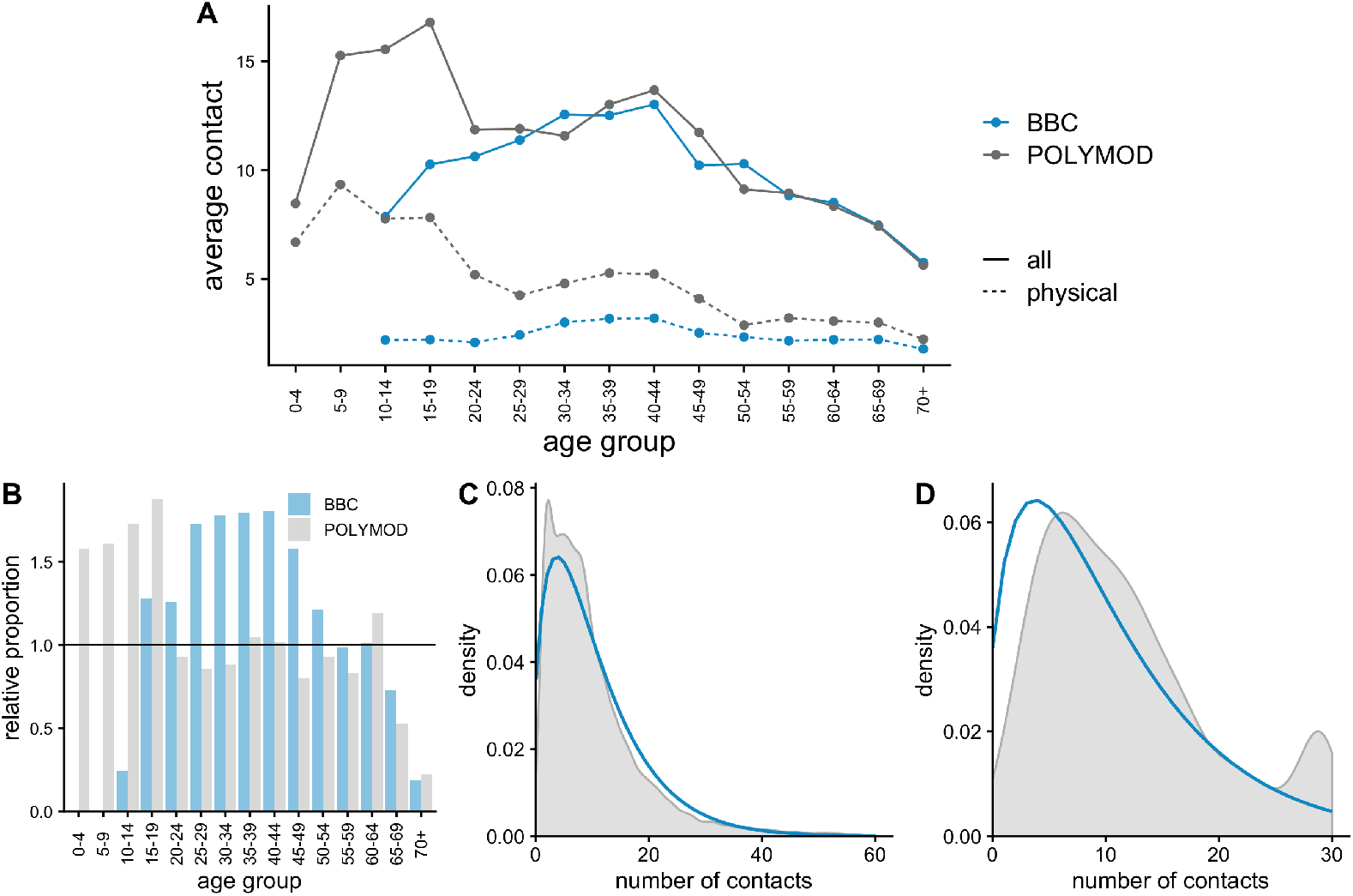
Comparison of BBC and POLYMOD datasets. (A) Mean number of contacts for average person (assuming 2018 population structure) (B) proportion of participant age-groups relative to UK population proportions (ONS 2018 data) in POLYMOD study (gray) and BBC study (blue). Distribution of the number of contacts per participant with its MLE-fit of negative-binomial distribution indicated in blue for (C) BBC data (size = 1.68, *µ* = 10.47), and (D) POLYMOD data (size = 2.98, *µ* = 11.81).

Out of all reported contacts in the POLYMOD dataset, 46.3% were physical (involved touch), 50.2% didn’t involve touch, and the rest were unspecified, resulting in comparable levels of reported conversational and physical contacts. In the BBC dataset, participants reported about three times more conversational contacts than physical (75.6% of reported contacts were conversational and 24.4% were physical). Compared to the general population, POLYMOD oversampled younger groups by design (i.e. ages under 20) and the BBC data ended up oversampling adults (in particular ages 25-49); both studies undersample ages over 65 (Fig 6A). The distribution of the number of contacts reported across all participants followed a negative-binomial distribution with mean 10.47 for BBC data and 11.81 for POLYMOD data (Fig 6C and D, respectively). The POLYMOD data was right censored at 29 contacts, which is evident in the density plot (Fig 6D).

### 5.1 Consequences for disease dynamics

As the contact matrices are related to next generation matrices, the differences in structure between the POLYMOD and BBC matrices have direct consequences for disease dynamics. Here, we are particularly interested in the age-groups that are most responsible for transmission, which is described by the dominant eigenvector of the next generation matrix. We compare POLYMOD matrices for Great Britain consisting of the average number of contacts recorded per day per survey participant that are available from Mossong et al. [22] Table S5. Even though the raw data from POLYMOD is available, these matrices are most commonly used, which is why we choose them for the comparison with BBC matrices. Ages are grouped in 5-year bins until the age of 70, and a single 70+ age group (total of 15 age-groups). For the comparison of matrices we follow the same grouping (Fig 7), and make both BBC and POLYMOD matrices reciprocal using the 2018 population vector for England and Wales [1].

**Figure 7:**
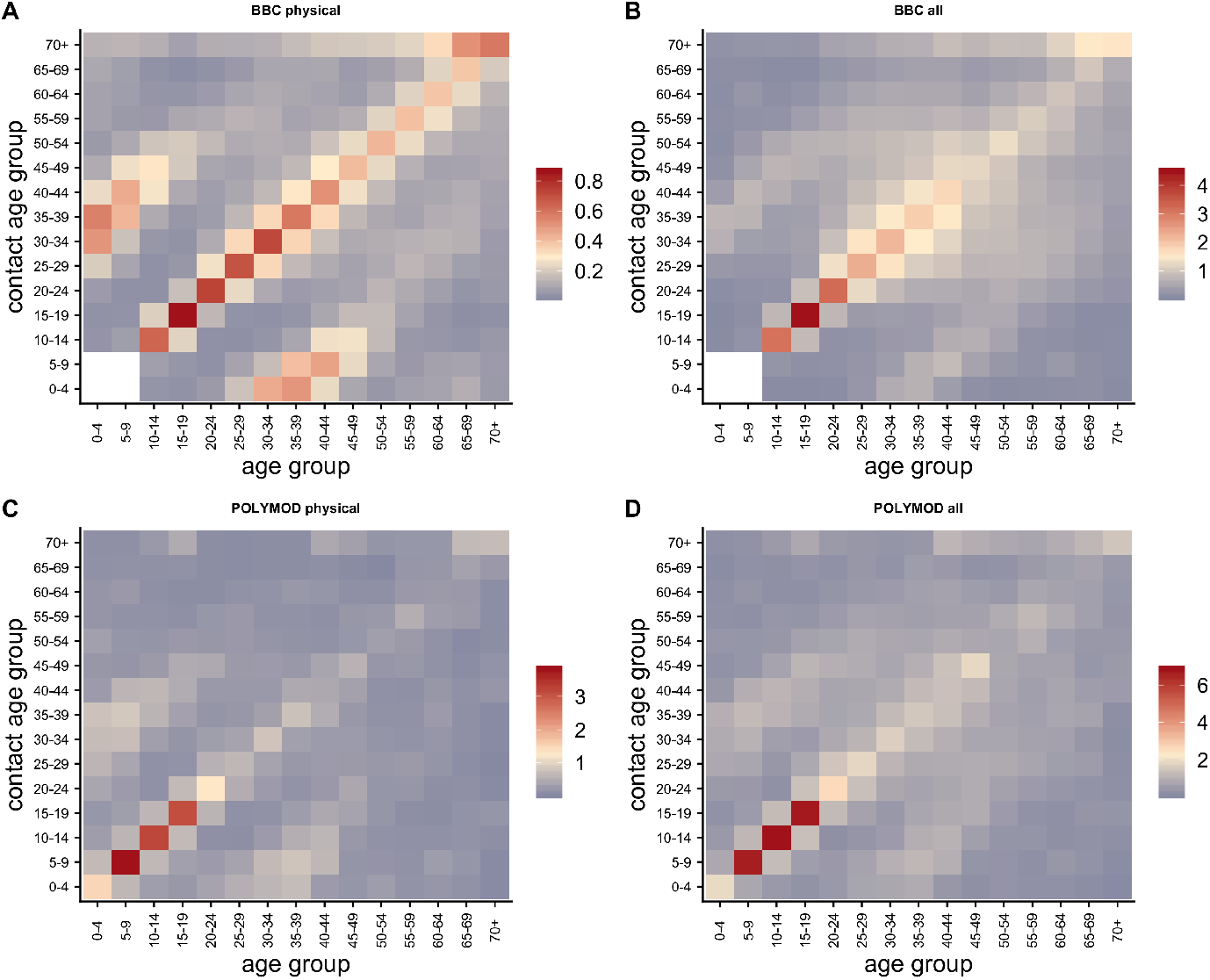
Comparison of BBC and POLYMOD mixing matrices for physical contact only and all contact. (A) BBC physcial contacts. (B) BBC all contacts. (C) POLYMOD physical contacts. (D) POLYMOD all contacts.

We address the missing data in BBC matrices in two ways: (1) we take the ages 13 and 14 for which we have data to be representative of the entire age-group 10-14, and (2) we fill out the information for the age-groups 0-4 and 5-9 from POLYMOD by scaling the missing square with the ratio of dominant eigenvalues of the symmetric subset of the BBC matrix without missing values, and the same subset of the POLYMOD matrix. The scaling factor *q* = *ρ*(BBC_*S*_)*/ρ*(POLYMOD_*S*_) where *S* designates this symmetric subset of the matrix and *ρ*() is the dominant eigenvalue (spectral radius) ensures that the dominant eigenvalue of the filled in BBC matrix stays intact.

The largest eigenvalue of the reciprocal contact matrix is the average number of different people contacted during one day by someone who has just been contacted [29] (this value is proportional to *R*_0_). The relevant type of contact for transmission (physical or all contact) will depend on the type of pathogen. For pathogens that require close sustained contact for transmission (such as commensal skin colonisers associated with nosocomial infections like *Staphilococcus aureous* [12]), physical contacts might be more relevant. For very easily transmissible pathogens like measles, some combination of physical and conversational contacts would better represent transmission. Figure 8A illustrates the difference between the average number of people contacted for a whole range of matrices ranging from physical contacts only to all contacts (by gradually adding conversational contacts in increments of 0.1). The average number of physical contacts in the POLYMOD dataset are more than twice that of the BBC dataset. However on average participants in the BBC dataset speak to two more persons a day than POLYMOD participants (7.9 compared to 5.9).

**Figure 8:**
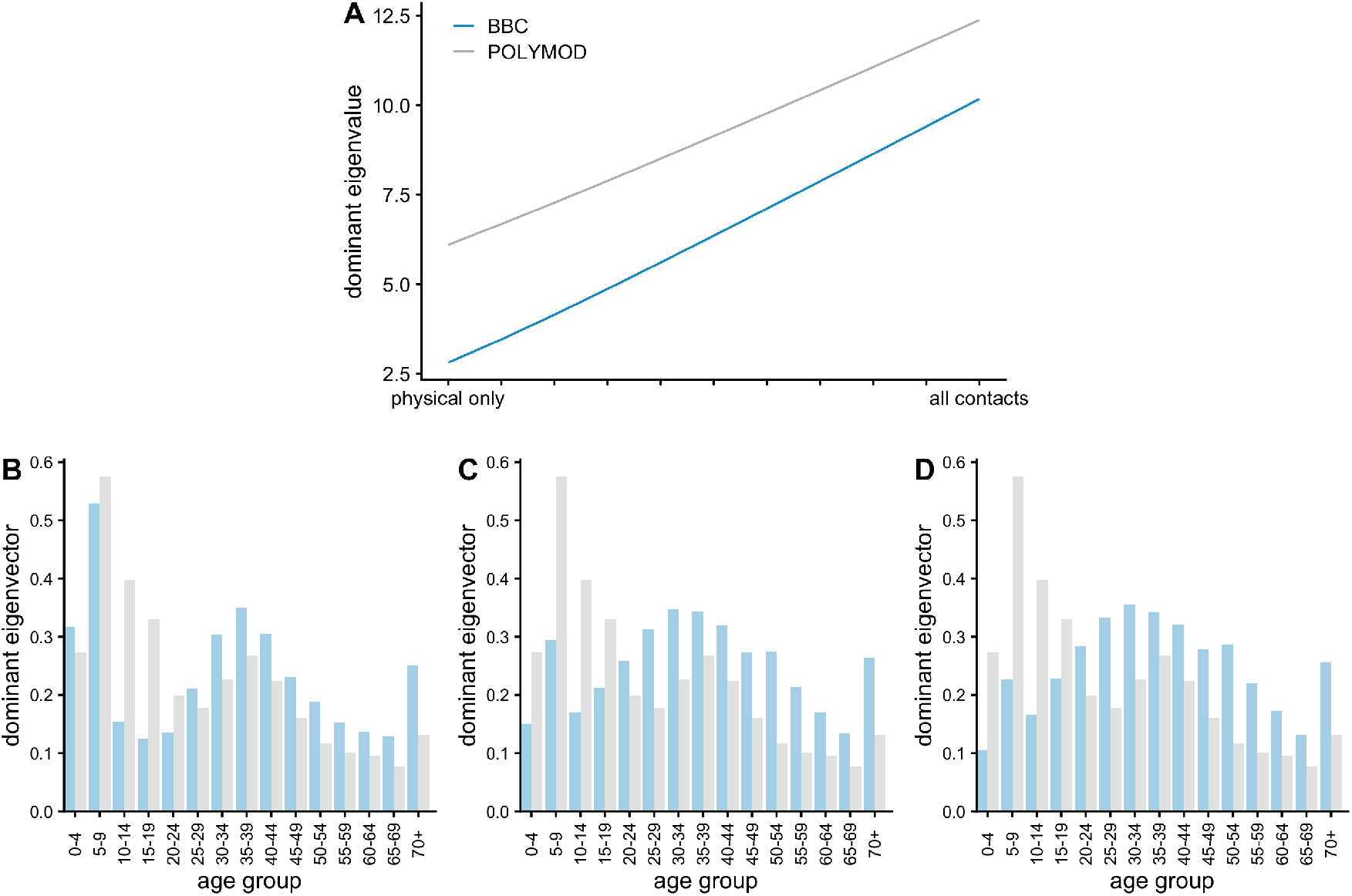
Comparison of BBC and POLYMOD mixing matrices. (A) Dominant eigenvalues for the resulting mixing matrices for POLYMOD (gray) and BBC (blue) as the contribution of conversational contacts increases from 0 to 1 (going from physical only contacts, to all contacts). Comparison of dominant eigenvectors for the BBC (blue) and POLYMOD (gray) mixing matrices for (B) physical only contacts (C) physical only plus half the conversational contacts, (D) all contacts.

The age-specific contributions to those overall contacts (or overall transmission during the early stages of an outbreak) are given by the magnitude of the dominant eigenvector. Fig 8B-D shows the relative contribution of different age-groups to overall transmission for physical contacts only (B), physical plus scaled conversational contacts by a half (C) and all contacts (D) for both the BBC and POLYMOD studies. Except for physical contacts, using BBC mixing matrices generally leads to more transmission in adult age-groups (particularly in ages over 25) whereas with POLYMOD dataset school-children are largely responsible for transmission regardless of how we construct the overall matrix.

## 6 Discussion

The BBC Pandemic study has the potential to provide extremely detailed insights into patterns of movements and social mixing in the UK, which will be valuable for understanding the dynamics of circulating infectious diseases as well as informing the prevention and control of new outbreaks. Analysis of the full study dataset is ongoing to ensure that information relevant for epidemiological research can be made widely available, while also protecting participant privacy and anonymity. However, the emergence of the novel coronavirus disease COVID-19 [15] has created an urgent need for the best possible social mixing data to be made available to support the outbreak response, as well as for the possibility to use multiple sources of social mixing data to evaluate the uncertainty that stems from social mixing in evaluating public health interventions. It is therefore our hope that this detailed contemporary picture of age-specific mixing patterns will be of value to those modelling COVID-19 to improve the evidence base for decisions on potential control measures in the UK, as well as suggesting broader insights into social mixing that may be relevant to other countries as well.

The matrices we present can be directly incorporated into mathematical models of transmission to predict the dynamics of infection with and between key demographic groups and settings [18, 23, 2, 17]. The large scale nature of the BBC data presented, with over 36,000 users and over 378,000 contacts, made it possible to generate fine-scale age-specific social contact matrices across different contexts, and by week and weekend. These could be used to explore the impact of different intervention strategies that rely on social distancing to reduce the amount of mixing in the population (such as school closures, and working from home) on flattening and postponing the peak of an outbreak. There were some notable differences between these matrices and those presented in the POLYMOD study, which surveyed 1,012 people in Great Britain. In particular, POLYMOD participants report higher overall mean contacts than BBC participants (11.7 compared to 10.5 people on average), although this difference is reduced when both dataset are calibrated to the same population structure. More than a decade has passed since the POLY-MOD study, and it is possible that the average number of contacts in the UK may have dropped. This is particularly evident in the reduction of teenage contacts, which could be the signature of a real change in how teenagers interact with one-another and their preferred way of interacting with their friends shifting from face-to-face to social media over last several years [24]. While the mean number of conversational contacts reported by participants in the BBC study was higher than in POLYMOD (7.9 and 5.9 respectively), the mean number of physical contacts was significantly lower, and mostly limited to contacts at home. Moreover, POLYMOD by design oversampled children, whereas the BBC data oversampled adults, and hence may have captured more of the tail of the contact distribution in older age groups.

Incorporating social mixing patterns in different contexts and at different times of the week (weekend vs weekday) into mathematical models, it is possible to evaluate the potential effectiveness of a range of control measures targeting respiratory infections, including school closures [4, 14] and social distancing [11]. However, the precise combination of setting and type of contact that will be important for transmission will depend on the infection being considered. There is evidence that both physical contacts [18, 6] and conversational contacts [8] may be relevant for capturing the transmission dynamics of acute respiratory infections such as Streptococcus pneumoniae and influenza A/H1N1p, and for influenza, there can be substantial transmission in households [10] and schools [21, 5]. How to weight the respective contributions of conversational and physical contacts to overall population transmission will depend on the pathogen.

There are some additional limitations to the dataset presented here. First, children under the age of 13 were excluded for ethical reasons, which meant there was a gap in the matrices for participants in this group. Given the role of school-age children in transmission of many respiratory infections, we are missing important information on mixing in school-aged children. For a flu-like pathogen, this core group will be responsible for driving transmission in the wider population [2] which can be seen from indirect effects observed in other age-groups by targeting the vaccination of children [13]. In our previous work [17] we filled the missing square sub-matrix (the dimensions of it will depend on the size of the age-groups chosen in the model) after making contacts reciprocal with appropriate values from POLYMOD, and here we take an additional step of rescaling the missing sub-matrix so that the overall dominant eigenvalue of the matrix doesn’t change. The missing data could also be interpolated from surrounding regions and assuming the log-binomial distribution of contacts.

There are other possible biases as well - the day that the participants took part was not randomly assigned. Instead, the participants could choose themselves when to run the app which might have biased some to choose a particularly ‘interesting’ day when they are going to meet a lot of people, or travel somewhere unusual. In addition, the participants themselves were not sampled at random from the population but instead chose to take part. How they heard about the study might might have varied from whether they were reached through social media in the drive to recruit participants before broadcast, or consequent to watching the BBC4 programme, or through hearing about it from friends – all of which could lead to selection bias. Given the big social media exposure around the citizen science project, our participants were recruited largely in two time periods: in October 2017 after the launch of the app, and in March 2018 after the airing of the documentary ‘Contagion! The BBC4 Pandemic’, but the uneven recruitment of participants over time should not have much impact. There is evidence, at least among school-aged children, that social contact structure during term-time is relatively consistent over a period of several months [19]. Contacts can also change between term-time and school-holidays [8] and with the health status of participants; individuals typically make fewer social contacts when they have ILI compared to a normal day [28]]. It may therefore be necessary to combine the matrices presented here with other datasets to fully explore transmission dynamics over long periods and account for changes in behaviour according to health status.

Finally, by comparing the BBC mixing matrices to ones from POLYMOD we show that there are important differences in age-specific contributions to transmission with school-children driving transmission in POLYMOD, while in the BBC dataset adults are more responsible. The exception here is if mixing is driven by purely physical contacts when ages 5 to 9 are most responsible for transmission. These results have strong implications for control strategies (such as informing school closures) and using different underlying mixing patterns could lead to different policy recommendations. This illustrates the importance of using several sources of data for informing the age-specific mixing of the population to account for the uncertainty that stems from population mixing.

## Data Availability

Data in this manuscript is available in the form of contact matrices in XLS files.

## 7 Acknowledgements

AJK was supported by a Sir Henry Dale Fellowship jointly funded by the Wellcome Trust and the Royal Society (grant Number 206250/Z/17/Z). MLT was supported by the UK Engineering and Physical Sciences Research Council (EPSRC), grant number EP/N509620/1. We are grateful to Edwin van Leeuwen for helpful discussions on POLYMOD matrices. We would like to thank 360 Production, and in particular Danielle Peck and Cressida Kinnear, for helping to make the collection of this dataset possible, and all our study participants for giving up their time to contribute to this public science project. We are grateful to Anne Alexander and Hugo Leal for helpful discussions regarding the ethics considerations and data privacy.

---

**Figure S1:**
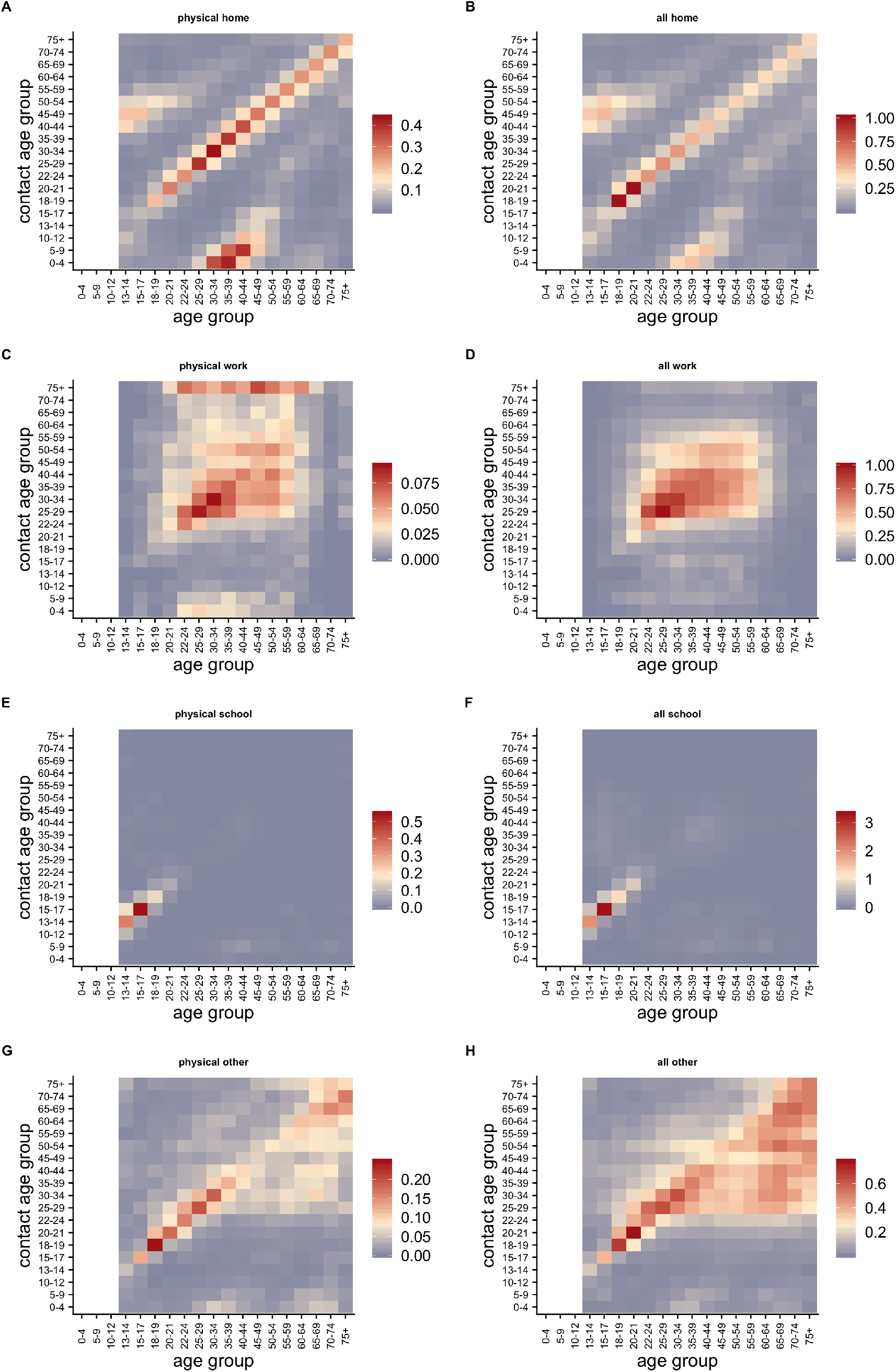
Raw contact matrices. Mean number of contacts reported by participants of given age groups, **M**, before accounting for reciprocity of contacts. Matrices are by physical contact only and by all contact (conversational and physical), in respective columns) and by different encounter context (home, work, school or other in respective rows). A) physical home, B) all home, C) physical work, D) all work, E) physical school, F) all school, G) physical other, H) all other.

**Figure S2:**
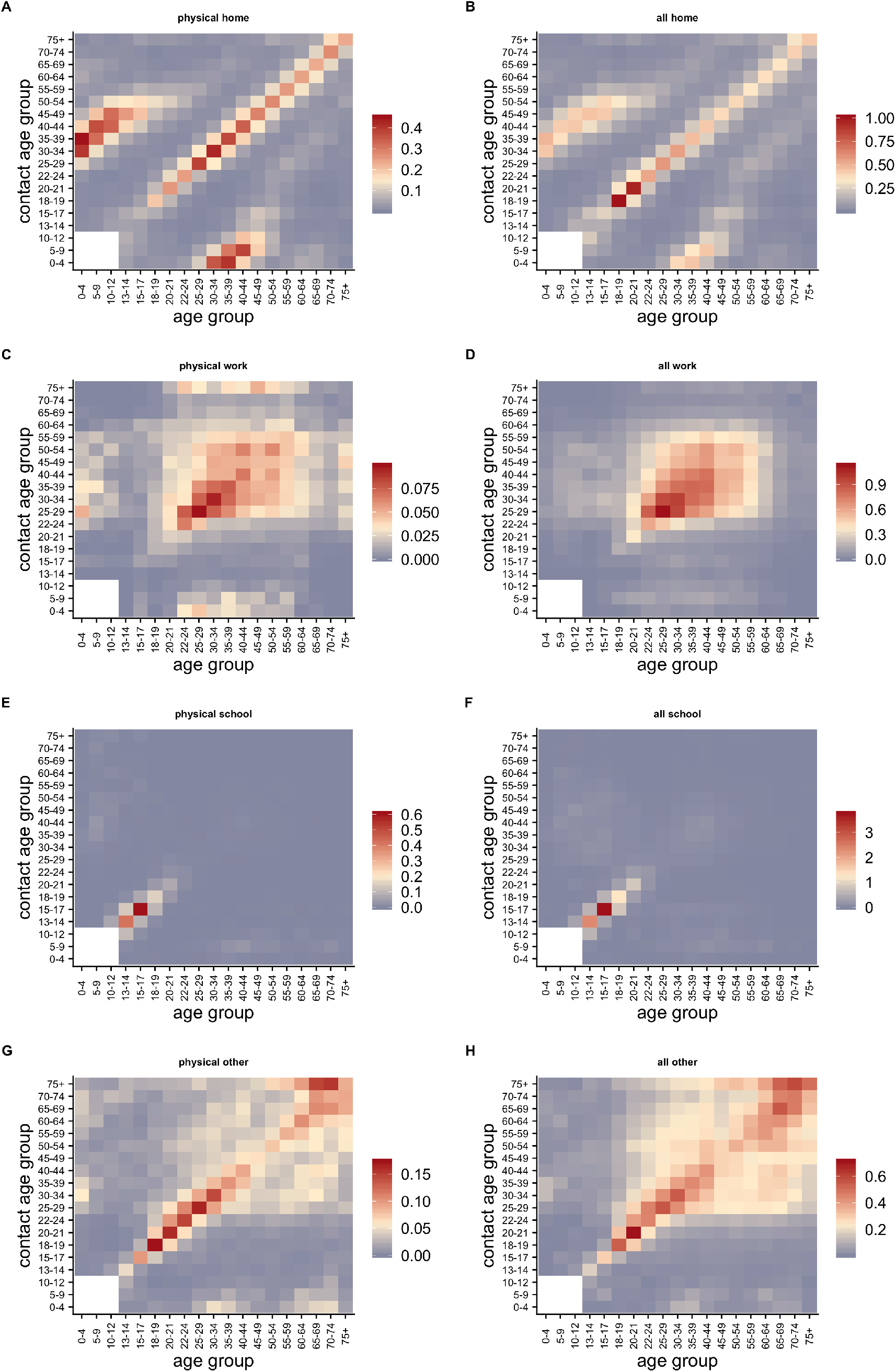
Contact matrices during the week adjusted for reciprocal contacts. Matrices are by physical contact only and by all contact (conversational and physical), in respective columns) and by different encounter context (home, work, school or other in respective rows). A) physical home, B) all home, C) physical work, D) all work, E) physical school, F) all school, G) physical other, H) all other.

**Figure S3:**
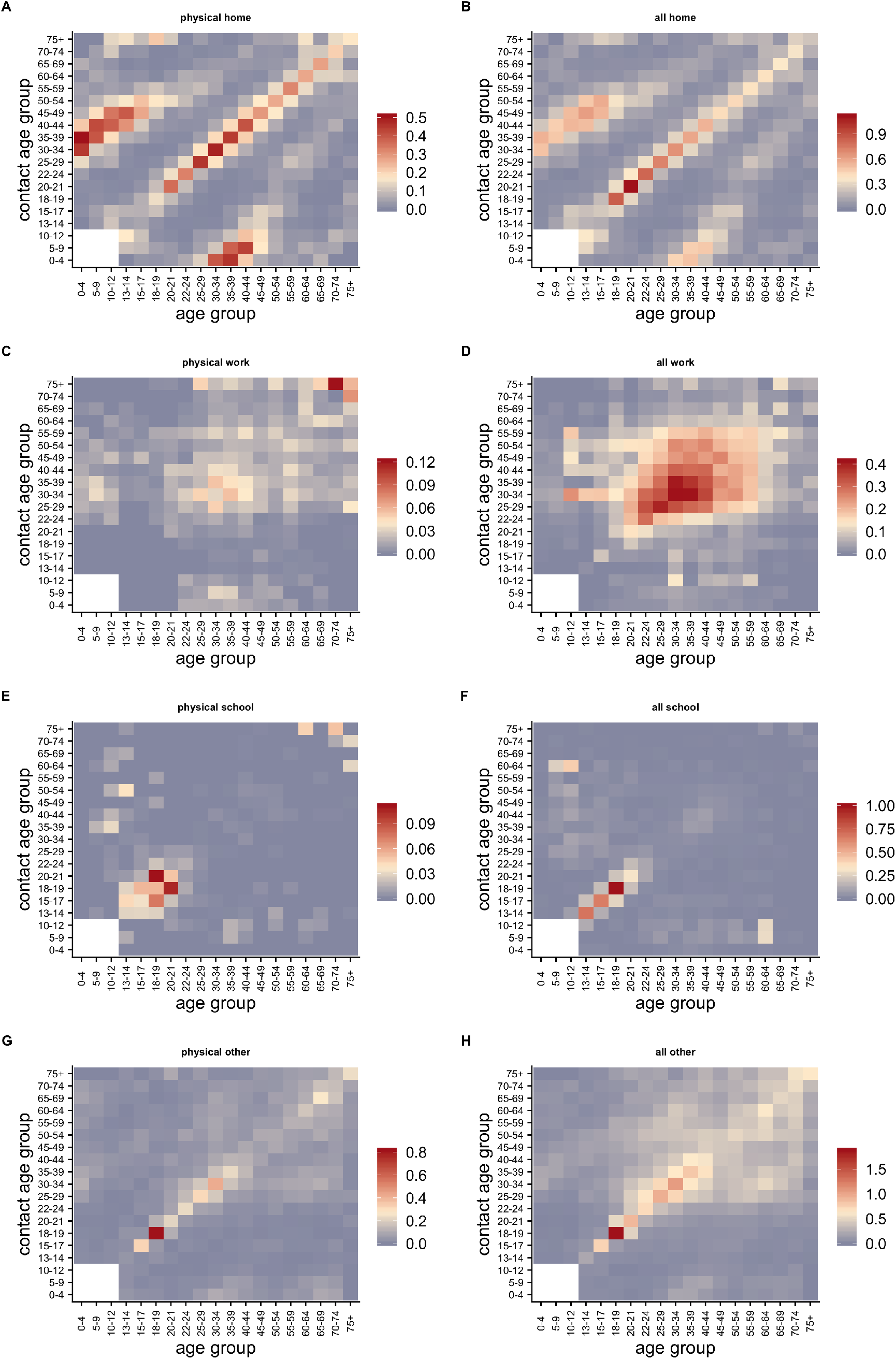
Contact matrices during the weekend adjusted for reciprocal contacts. Matrices are by physical contact only and by all contact (conversational and physical), in respective columns) and by different encounter context (home, work, school or other in respective rows). A) physical home, B) all home, C) physical work, D) all work, E) physical school, F) all school, G) physical other, H) all other.

